# Scaling SARS-CoV-2 Wastewater Concentrations to Population Estimates of Infection

**DOI:** 10.1101/2021.07.15.21260583

**Authors:** Edward H. Kaplan, Alessandro Zulli, Marcela Sanchez, Jordan Peccia

## Abstract

Monitoring the progression of SARS‐CoV‐2 outbreaks requires accurate estimates of infection rates. Estimation methods based on observed cases are biased due to changes in testing over time. Here we report an approach based upon scaling daily concentrations of SARS‐CoV‐2 RNA in wastewater to infections that produces representative estimates due to the consistent population contribution of fecal material to the sewage collection system.

---

Estimating the fraction of individuals infected in coronavirus outbreaks is of first‐order importance in monitoring epidemic progress and evaluating interventions meant to slow transmission. Given the lack of repeated representative COVID‐19 testing over time, researchers have attempted to estimate SARS‐CoV‐2 incidence from lagging indicators of infection including clinically diagnosed cases, hospitalizations, and deaths^1,2,3,4,5^. Even if the lags from infection until the occurrence of diagnosed COVID‐19 cases, hospitalizations, or deaths can be estimated over time, such indicators still depend upon time‐varying and non‐representative observations due to temporal changes in the rates and targeting of COVID‐19 testing, hospital admission and treatment policies, and undercounting of deaths from COVID‐19.

Recognizing these difficulties, we initiated daily sampling at a wastewater treatment plant (WWTP) serving a mid‐sized US municipality with the goal of obtaining a representative view of local coronavirus outbreaks. Nucleic acid was extracted from primary sewage sludge and the concentration of SARS‐CoV‐2 RNA was determined. Such RNA concentrations provide a proportional measure of the extent of infection in the community given the population’s consistent discharge of fecal material into the local sewage collection system. We previously reported daily SARS‐CoV‐2 wastewater concentrations during the March 2020 epidemic wave in this community^6^, and via an epidemic model with an assumed SARS‐CoV‐2 shedding distribution, reported the associated reproductive number *R*_*0*_ and cumulative incidence of infection^7^.

Based upon this previous work, we implement a simple model enabling estimation of the fraction of the population infected over time directly from the RNA concentrations in sewage sludge. The incidence of infection is determined from the start of the pandemic in Connecticut USA (March 19,2020) through May 31, 2021, and results are compared to three independently developed estimates based on cases, hospitalizations, and deaths.

Figure 1a reports the total and positive daily number of COVID‐19 tests conducted on the residents of the four towns served by the WWTP. These data demonstrate the large increase in testing as the pandemic progressed, while the positive test results illustrate the two major COVID‐19 waves experienced in the area with the second wave yielding higher daily case rates over a longer duration compared to the first. Figure 1b plots SARS‐CoV‐2 RNA concentration measured in sewage sludge over this same time period. This figure also shows two waves of infection. Unlike the number of positive tests, note that for the wastewater data the first wave, though shorter in duration, peaks at concentrations similar to the second wave. This demonstrates that the early lack of testing impacted the accuracy of reported COVID‐19 case information.

**Figure 1(a):**
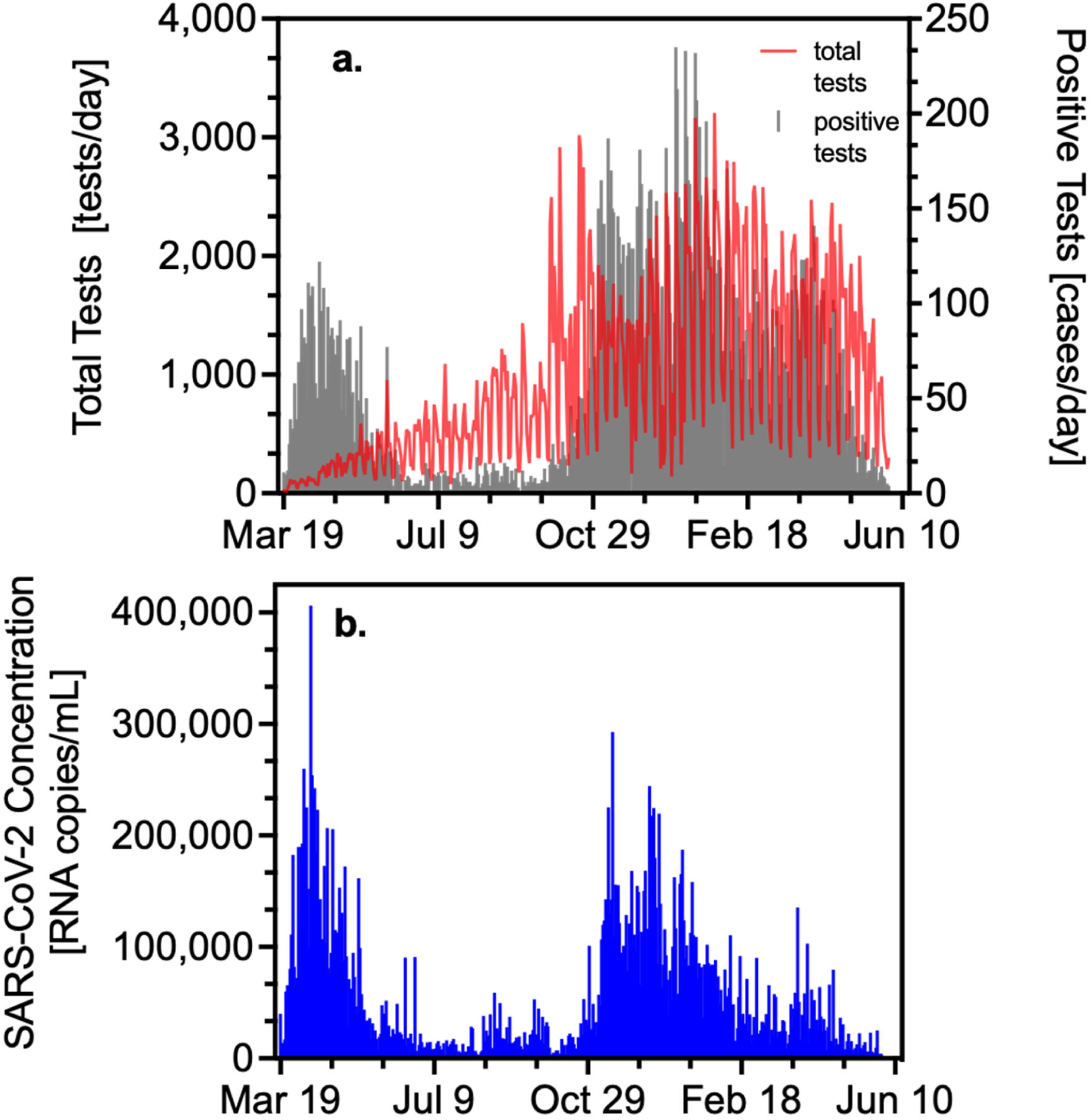
total and positive COVID‐19 tests over time. **(b)** SARS‐CoV‐2 RNA wastewater concentration (copies/mL sludge).

Applying equation (1) (see Methods) to the data contained in Figure 1b yields the cumulative fraction infected in the population over time (Figure 2). We estimate that 33.6% (95% CI [24.3%, 42.9%]) of the population was infected by May 22, 2021. Figure 2 also shows point estimates for the cumulative fraction of the population infected in New Haven County (which subsumes the treatment plant population) produced by three computationally intensive statistical models using completely different methods and data sources including COVID‐19 cases and deaths^3^; cases, deaths, hospitalizations, and close‐contact measures deduced from cell phone geolocation data^4^; and deaths alone^5^. These models demonstrate strikingly similar shapes to and bracket the results of the wastewater‐based estimates. Model 1 (ref. ^3^) hugs the wastewater model’s upper 95% confidence limit, Model 2 (ref. ^4^) hugs the wastewater model’s lower 95% confidence limit, and Model 3 (ref. ^5^) falls just beneath the point‐estimate trajectory of equation (1).

**Figure 2:**
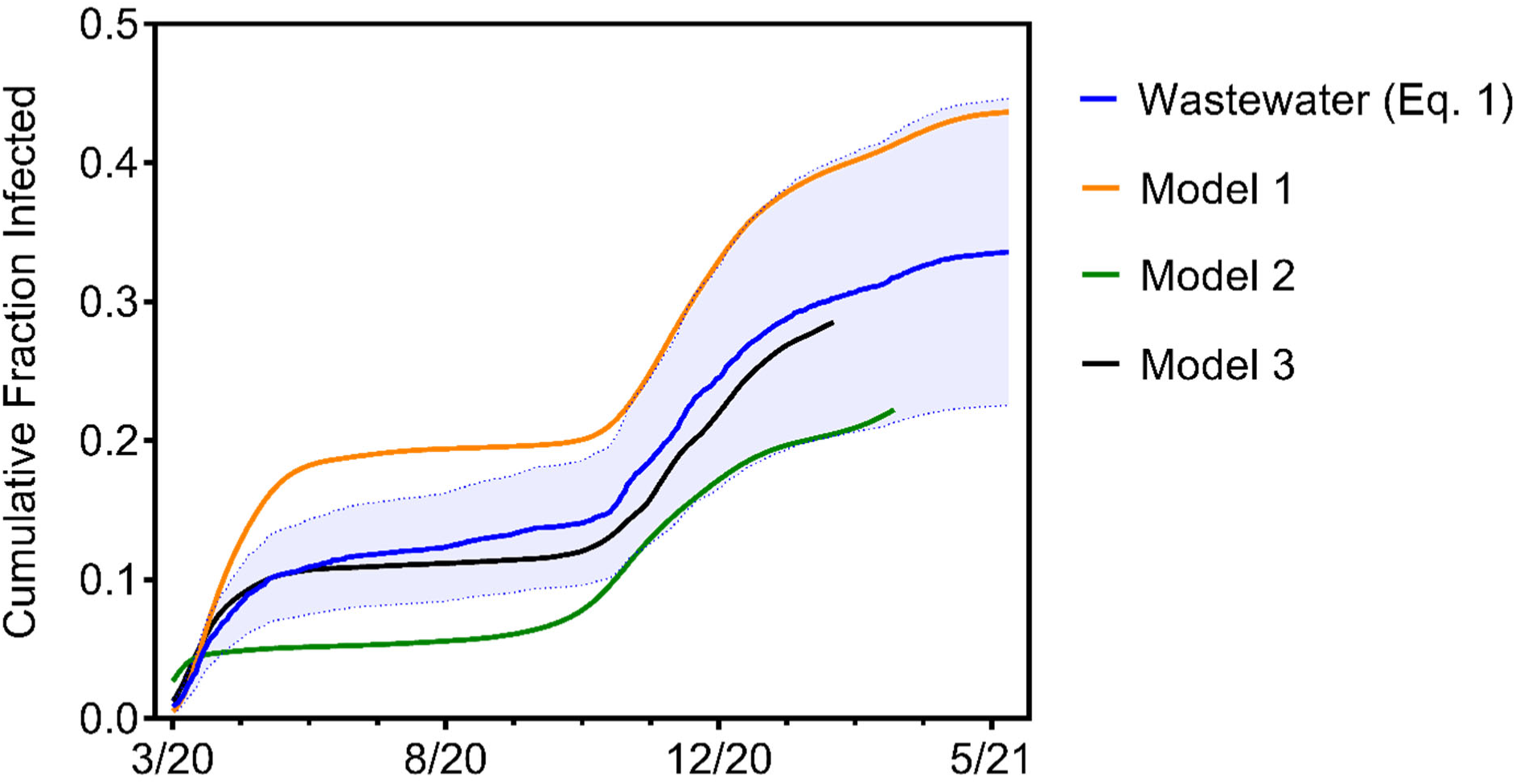
Estimated cumulative incidence of infection with 95% CIs in the population served by the WWTP based on the SARS‐CoV‐2 RNA concentrations shown in Figure 1b using equation (1). Also shown are cumulative incidence estimates for New Haven County produced by three different statistical models. While all four models show similar trajectories over time, the estimates from equation (1) are the middle of the range exhibited by the other models.

The utilization of wastewater‐based epidemiology surged during the COVID‐19 pandemic with applications to outbreak detection and tracking temporal trends^8,9,10^. This report presents a major advance for wastewater surveillance by using a simple scaling model to directly estimate the fraction of persons in a population infected over time from SARS‐CoV‐2 RNA concentrations in wastewater. This approach circumvents problems with non‐representative sampling inherent in models based on diagnosed cases, hospitalizations, or deaths, and in principle can be applied in any location where continuous wastewater sampling over time is possible.

## Methods

The number of total tests and confirmed and probable COVID‐19 cases was provided by the Connecticut Department of Public Health (CT DPH).

SARS‐CoV‐2 RNA concentrations were quantified in the primary sludge of the New Haven, CT, USA wastewater treatment plant, which serves 200,00 residents. Nucleic acid was extracted using commercial kits (Qiagen, RNeasy Powersoil Total RNA kit and Zymo, Quick‐RNA Fecal/Soil Microbe Microprep). Nucleic acids were measured by spectrophotometry, the concentration adjusted to 200 ng μL‐1 (NanoDrop, Thermo Fisher Scientific) and SARS‐CoV‐2 RNA concentrations were quantified through one‐step qRT‐PCR kit (BioRad iTaq™ Universal Probes One‐Step Kit) using SARS‐CoV‐2 N1 and N2 primer sets for quantification in accordance with previously described protocols^6,11^. Further details regarding the construction of the SARS‐CoV‐2 RNA concentration dataset appear in the Supplementary Information.

The concentration of SARS‐CoV‐2 RNA in sewage sludge lags the population incidence of SARS‐CoV‐2 in accord with the generation time distribution (also referred to as the shedding load distribution) from infection to transmission, the mean of which is approximately 9 days^7,12^. Letting π_*t*_ denote the fraction of the population that is newly infected on day *t* (the incidence of infection) and l denote the mean generation lag, the SARS‐CoV‐2 RNA concentration measured on day *t, Z*_*t*_, should approximately reflect the incidence of infection l days earlier, that is, *Z*_*t*_ ≈ *k*π_*t*‐l_ where *k* is the constant of proportionality, and consequently the cumulative fraction of the population infected by the end of day t, given by 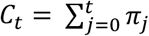, should approximately follow 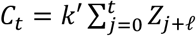 where *k*′=1/*k* is the constant of proportionality scaling SARS‐CoV‐2 RNA concentration to infections per person. Given the cumulative number of infections 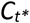 as of some particular date *t*\*, let 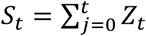 denote the cumulative sludge RNA observed through day *t*. Then the scaling constant k′ can be evaluated from the relation 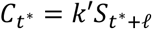 yielding 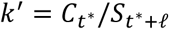. Substituting back into the equation for cumulative incidence up to an arbitrary day *t* yields 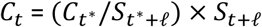. The cumulative incidence of infection in the 200,000 population served by the WWTP treatment plant was previously estimated as 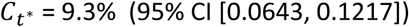 as of *t*^*\**^ = May 1, 2020 with a mean generation lag of 8.9 days which we round up to l = 9 (ref. ^7^). Substituting yields our scaling of cumulative RNA concentration to cumulative incidence shown in Figure 2 as

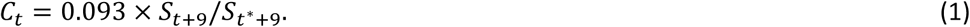

Confidence intervals follow from the variance of *C*_t_ estimated via the delta method^13^.

## Supporting information

Study dataset

## Data Availability

All data used in study included as supplemental information.

## Data Availability

All data employed in this report are available in the Excel file contained in the Source Data.

## Acknowledgements

We thank Stephen Bart from the Connecticut Department of Public Health for assembling the COVID‐ 19 testing data for the four towns served by the WWTP. We also thank Olga Morozova and Forrest W. Crawford for sharing the cumulative incidence estimates produced by their model for New Haven County.

## Supporting Information

### Extracting SARS‐CoV‐2 RNA from Wastewater and Scaling for Consistent RNA Measurement

From March 19, 2020 through September 30, 2020, well‐mixed primary sludge was added directly to Qiagen’s RNeasey PowerSoil Total RNA commercial extraction kit^1^. Supply problems required us to change the RNA extraction kit used to Zymo Quick‐RNA Fecal/Soil Microbe Microprep for samples collected from October 1, 2020 to May 31, 2021^2^. To assure consistency between measurements based on these two different RNA extraction kits, Zymo extractions were also performed on stored samples originally analyzed with the Qiagen kit corresponding to 66 of the 74 days between March 19, 2020 and May 31, 2020 inclusive (eight of the original 74 samples were fully utilized and re‐extraction with the Zymo kit was not possible). This enabled us to covert Qiagen RNA values to equivalent Zymo measurements as follows: letting *Q*_*t*_ and *Z*_*t*_ denote the SARS‐CoV‐2 RNA concentrations measured with the Qiagen and Zymo kits respectively on day *t* (*t* = 1, 2,…,66), we assumed that the Zymo RNA concentration on day *t* followed a normal distribution with mean *E*(*Z*_*t*_) = *a + bQ*_*t*_ and variance *Var*(*Z*_*t*_) = *cE*(*Z*_*t*_) with the variance‐proportional‐to‐mean formulation following from prior statistical analysis of SARS‐CoV‐2 RNA concentrations^3^. The parameters *a, b* and *c* were estimated by maximizing the log‐ likelihood function

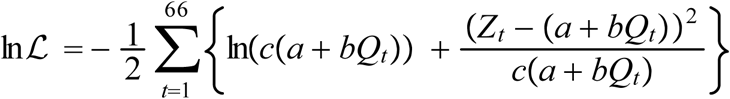

using the Analytic Solver in Excel^4^. The covariance matrix of the estimated parameters was computed following standard maximum likelihood theory by inverting the negative Hessian matrix of the log likelihood function^5^, and standard errors were extracted as the square root of the diagonal of the covariance matrix; these computations were also performed in Excel. The estimated intercept was not statistically different from zero (*â* = 9,180, 95% CI [‐6,870, 25,200]), so a model with no intercept (*a* = 0) was fit yielding 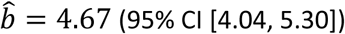 and *ĉ* 30,300 (95% CI [19,190, 41,400). The models with and without an intercept were further compared by computing twice the difference in the log likelihood values between the models yielding χ^2^ 2.2 (*p* = 0.14 at 1 degree of freedom, not significant). Models with and without an intercept assuming that *Var*(*Z*_*t*_) is constant, corresponding to ordinary least squares regression analysis, were also estimated but these models produced substantially lower log likelihood values compared to the models described above (with the same number of estimated parameters) indicating substantially worse statistical fit, and were thus not considered further.

### Assembling the Consistent Zymo RNA Concentration Dataset

We used the observed Zymo RNA concentrations together with the scaling model with no intercept described above to produce a consistent Zymo record of coronavirus RNA concentrations in the wastewater over the 439 days from March 19, 2020 to May 31, 2021. This was accomplished as follows:

1. On any day with an observed Zymo RNA concentration, that observed value was used. This accounts for 309 of the 439 days in the data.
2. On any day where a Qiagen measurement was made but no corresponding Zymo measurement was available, a Zymo value was estimated as of 4.67 times the associated Qiagen value based on the scaling model described above. This accounts for 125 of the 439 days in the data.
3. On any day missing both Qiagen and Zymo values (corresponding to the five days during the study period that the treatment plant did not provide a sample out of 439 days total), missing Qiagen values were interpolated from nearby observed values, and these interpolated values were then multiplied by 4.67 to produce an estimated Zymo value in accord with the scaling model described above. Specifically: Qiagen values missing on May 3 and May 6 of 2020 were interpolated by averaging the observed Qiagen values two days before and two days after the missing values, while Qiagen values missing on July 20, 21 and 22 of 2020 were interpolated linearly as 0.25, 0.5, and 0.75 times the distance respectively from the observed Qiagen value on July 19 to the observed value on July 23.

The consistent Zymo dataset appears in Dataset S1, and indicates which values were measured directly or scaled from Qiagen measurements (and for the latter which Qiagen values were observed versus interpolated and the means of interpolation).

### Source Data (separate file)

Date, Number of COVID‐19 Tests, Number of Positive COVID‐19 Tests, Qiagen RNA Concentration, Zymo RNA Concentration, Cumulative Zymo RNA Concentration, Wastewater SARS‐CoV‐2 Cumulative Incidence (Equation (1)), Upper 95% CI, Lower 95% CI, Estimated SARS‐CoV‐2 Cumulative Incidence from Models 1, 2 and 3

